# A Plagiarism Paperdemic - Plagiarism in infection journals in the era of COVID-19

**DOI:** 10.1101/2022.08.04.22278404

**Authors:** Rahma Menshawey, Esraa Menshawey, Ahmed Mitkees, Bilal A. Mahamud

## Abstract

**Background:** The COVID-19 pandemic has caused drastic changes in the publishing framework in order to quickly review and publish vital information during this public health emergency. The quality of the academic work being published may have been compromised. One area of concern is plagiarism, where the work of others is directly copied and represented as one’s own. The purpose of this study is to determine the presence of plagiarism in infection journals in papers relating to the COVID-19 pandemic.

**Methods:** Consecutively occurring original research or reviews relating to the COVID-19 pandemic, published in infection journals as ranked by SCOPUS Journal finder were collected. Each manuscript was optimized and uploaded to the Turnitin program. Similarity reports were then manually checked for true plagiarism within the text, where any sentence with more than 80% copying was deemed plagiarised.

**Results:** A total of 310 papers were analyzed in this cross-sectional study. Papers from a total of 23 journals among 4 quartiles were examined. Of the papers we examined, 41.6% were deemed plagiarised (n=129). Among the plagiarised papers, the average number of copied sentences was 5.42±9.18. The highest recorded similarity report was 60%, and the highest number of copied sentences was 85. Plagiarism was higher in papers published in the year 2020. The most problematic area in the manuscripts was the discussion section. Self plagiarism was identified in 31 papers. Average time to judge all manuscripts was 2.45±3.09. Among all the plagiarized papers 72% belonged to papers where the similarity report was ≤15% (n=93). Papers published from core anglosphere speaking countries were not associated with higher rates of plagiarism. No significant differences were found with regards to plagiarism events among the quartiles.

**Conclusion:** Plagiarism is prevalent in COVID19 related publications in infection journals among various quartiles. It is not enough to rely only on similarity reports. Such reports must be accompanied by manual curation of the results with an appropriate threshold to be able to appropriately determine if plagiarism is occurring. The majority of plagiarism is occurring in reports of less than 15% similarity, and this is a blind spot. Incorporating a manual judge could save future time in avoiding retractions and improving the quality of papers in these journals.

“A poor original is better than a good imitation.”

**— Ella Wheeler Wilcox**

## Intro

COVID-19 is caused by the SARS-COV-2 virus which was first discovered in Wuhan China in 2019(He et al. 2020). In March 2020, the World Health Organization (WHO) declared it a pandemic. Since then, research on the SARS-CoV-2 virus has taken the scientific world by storm, amassing more than 24 000 preprints to date. In an effort to overcome scientific publishing barriers, publishers and journals have adopted a fast-tracked means to publish COVID papers(Carvalho et al. 2021). The development of preprint servers has re-vamped the scientific publishing model, whereby papers on COVID-19 can be published without peer-review, and are publicly accessible(Añazco et al. 2021a). While all these changes are welcome given the seriousness of this pandemic, it may have inadvertently compromised the integrity and quality of the papers being published on this topic. Evidence of compromised academic integrity can be seen in the number of retracted COVID-19 papers as listed by Retractionwatch.com (238 listed articles as of 2022-06-28). The COVID-19 pandemic has tested scientific integrity on an international scale, with many complaints of fraudulent science and lack of academic ethics in order to quickly publish a paper on this popular topic. These actions are not without consequence and have resulted in the financial loss, academic punishments, retractions and more(Dinis-Oliveira 2020).

One aspect of academic integrity that may have been compromised and has yet to be studied is plagiarism. Plagiarism is defined as the appropriation of another person’s work (Aronson 2007). Within the scientific academic construct this may constitute blatant copying of work that is not one’s own, in the form of written work or ideas. IN publications, it may be observed as direct copying of text from another paper, or excessive copying of one’s own previously published work known as self-plagiarism(Burdine et al. 2018). Most journals have policies denouncing plagiarism and use a program to check for similarity before a manuscript reaches reviewers. Examples of these programs include Turnitin, and iThenticate(Meo and Talha 2019). It is important to stress that similarity and plagiarism are very contrasting issues. Similarity maybe inevitable in some specialties, in areas of the manuscript such as the methods where the technical language is redundant or offers no alternative (consider the phrase: P values less than 0.05 were considered significant). These programs provide a similarity report, which is not a definitive indicator of plagiarism (Meo and Talha 2019). To elaborate, a high similarity report may likely denote plagiarism, however, most journals have a threshold of acceptable level of similarity. This number may range between 15-18%, and maybe lower in higher ranking journals. A blind spot to plagiarism appears here; a low level of similarity does not automatically exclude plagiarism, as plagiarism can be directly copied text well within the given range of the limit selected by the journal. Another blind spot can occur in the review process, where it is unclear if journals employ a second re-check for similarity after suggestions have been sent by reviewers to the author. At that moment, changes to the manuscript may have introduced added similarly, and even plagiarism to the text. It is for this reason that a manual check, that comprises human judge evaluation of the similarity report, would be needed to truly determine if plagiarism exists. The aim of this study was to evaluate the presence of plagiarism in COVID-19 related papers published in a variety of infectious disease journals, in a manual way using the similarity report provided by the Turnitin program.

## Methods

A total of 310 consecutively retrieved manuscripts were examined in this cross-sectional study. The present study did not involve any human subject participation, and the entire analysis was performed using the commercially available plagiarism detection software Turnitin. Hence, it is exempted from the Institutional Review Board (IRB) review and approval.

We surveyed papers published in infection journals of various quartiles as listed on SCOPUS journal rankings for original research and reviews relating to the COVID-19 pandemic from Jan 1 2020 to Dec 31 2021. The first 5 journals from each quartile were selected after an initial screen of the journal title and scope to ensure that the journal focused on a general scope of infectious diseases. As an example, if a journal with the title of “Bilharziasis Monthly”, as its scope did not cover the topic of COVID-19 and or it did not have an adequate or substantial number of published research or review papers on the focus of COVID-19. A list of all the examined journals and their quartiles is available in table 1. Five journals were selected from each quartile with the exception of quartile 4 which required the use of 8 journals to prevent significant differences from occurring in this quartile as a result of small sample size.

**Table.**
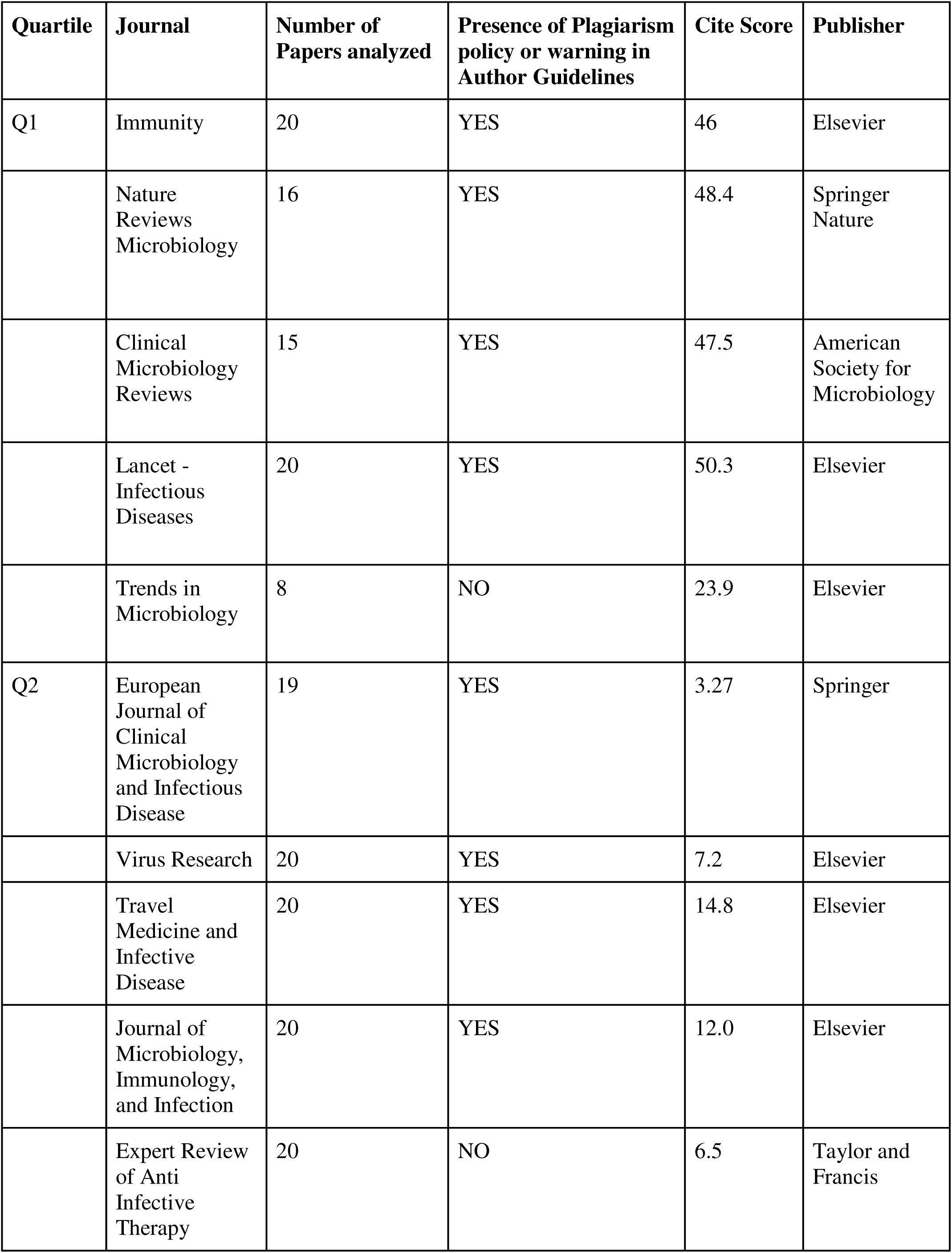
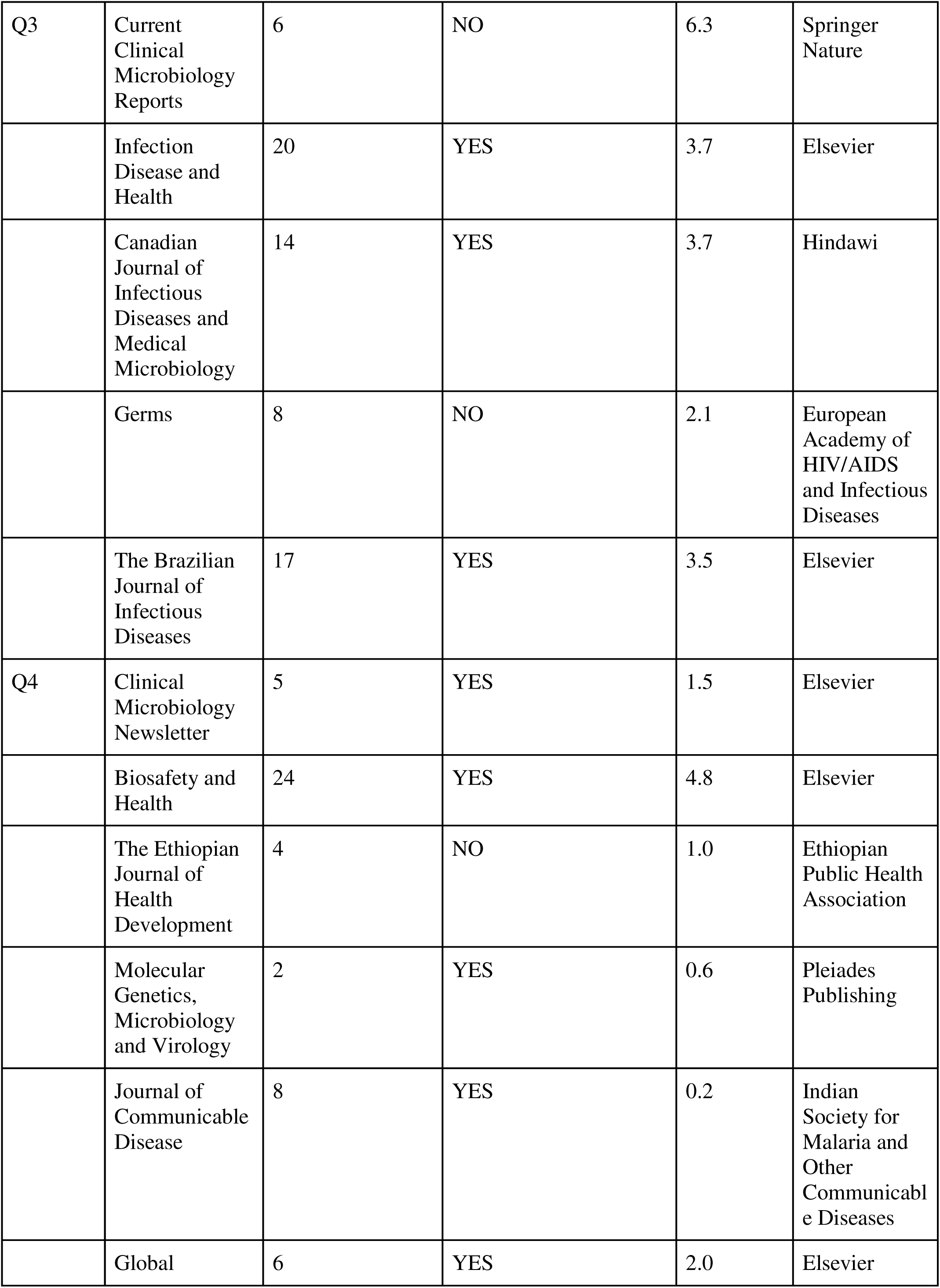
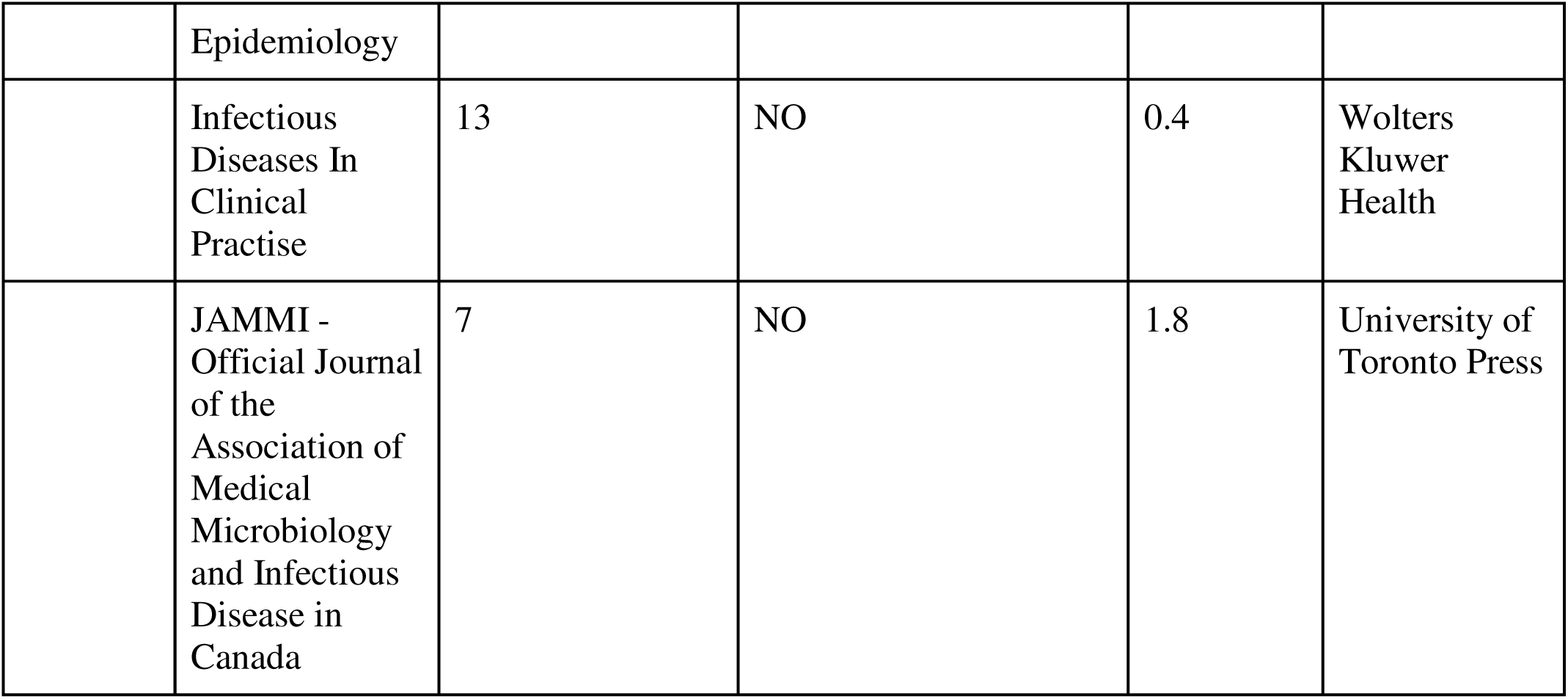
Journals Examined and Presence of Plagiarism Warning.

Papers were selected through each journal’s search engine in their order of publication using key words including; “SARS-COV-2”, and “COVID-19”. We excluded from our search the following; commentary, case reports, case reviews, editorials, letters to the editors, highlights, abstracts only, etc, (any non original research or review paper). We also excluded any non-English article. We only included papers for which the full-text was available (and as this pandemic was a public health emergency all papers full text were available). Papers met our initial screening via title, abstract and further reading to ensure that the selected papers were indeed relevant to the COVID-19 topic. All eligible papers were selected, and an optimized manuscript was extracted from each paper that met our inclusion criteria. As all COVID-19 related papers were made publicly accessible by journals and publishers, all the papers assessed in our study were accessible in their full text.

### Optimized Manuscripts

Optimized manuscripts were developed for each of the papers that met our inclusion criteria. This idea was inspired by the methods of Higgins et al(Higgins et al. 2016). Our optimized manuscripts included only the abstract, introduction, results, discussion and conclusion of each paper. We did not include the following; key words, methods, tables and figures with their corresponding legends, references, acknowledgments, conflict of interest statements, and any other part of the manuscript not directly related to the scientific content of the article. These optimized manuscripts were then uploaded to the Turnitin program and analysed.

The software used in this study is the Turnitin program. Turnitin is an internet-based tool developed by iParadigms LLC for the purpose of recognizing similarity among electronically submitted documents, and is often used by institutions to detect plagiarism. Its database provides a substantial repository of over 70 billion webpages, 1 billion student papers, and scholarly content of over 1700 publishers(SA and Talha 2019).

A total of 310 papers met our inclusion criteria, for which optimized manuscripts were developed based on our described methods (see table 1).

The following settings and exclusions were applied to the Turnitin program for each uploaded optimized manuscript:

- Under filter and settings: exclude quotes, exclude bibliography, exclude sources that are less than 10 words.
- Only periodicals, journals, publications

The following was excluded from the results of the similarity report:

- The original source being analysed. This included any other external link ie a website or repository, that was hosting this exact manuscript [examples of websites included - pubmed.com, x-mol.com]. Furthermore, with the introduction of preprint service for COVID19 related papers, if the exact manuscript (as identified by a combination of similar title, authors and topic) was identified by the Turnitin program, it was excluded.
- Publications that were published after the date of the analysed article. For example, if an article was published Jan 1st 2020, and the Turnitin program matched it in similarity with another article published Jan 1st 2021, it was excluded from the similarity report. Of note, due to the majority fast tracked peer review for COVID-19 related papers, we excluded papers that were matched up to within 1 month before the analysed paper. For example, if the article being analyzed was published Jan 1st 2021, and a match was made with an article published December 1st 2020, it was excluded from the results of the similarity report.

The final report was downloaded and analysed for results.

### Outcomes

Our outcomes included

- The results of the similarity report as determined with our filters, settings, and exclusion criteria
- Country of origin, as determined by the country of the corresponding author
- Language of country of origin as determined through a Google search.
- Number of authors as determined by the number of authors listed for each paper
- Whether true plagiarism was identified in the manuscript through manual review of the similarity report in comparison to a defined set of rules.
- Number of plagiarised sentences and where they were located (abstract, introduction, results, or discussion and conclusion)
- The area containing the greatest number of plagiarised sentences
- The presence of self-plagiarism. This was determined by if the majority of the similarity was coming from any other published paper belonging to any of the listed authors, that was also the top match identified by the Turnitin program.
- Time in minutes that it took to manually review the similarity report

### Determination of Plagiarism and Results

Each optimized manuscript similarity report, that was subjected to a Turnitin screening, was then analysed for plagiarism manually.

Our criteria for plagiarism was the following;

- If 80% of a sentence was found to be identical to a previously published sentence from any other paper. This was manually calculated and determined. Similarity on the Turnitin program is identified through highlighting. Each highlighted word was counted and divided by the total number of words in that sentence. If our threshold of 80% was met, then this sentence was counted as a plagiarised sentence. If a sentence was in direct quotations, it was not included (i.e. filtered through the Turnitin program).
- If a single sentence was determined to be plagiarised, then the whole manuscript was scored as plagiarized.
- The number of plagiarized sentences in each section (designated as abstract, introduction, results, and discussion/conclusion) was tallied.

### Exclusion criteria for plagiarism

We excluded the following sentences from being tallied as part of plagiarised text:

- Standard sentences - sentences that are descriptive, or definitions were excluded, and or were determined through judges (the authors) that they could not be worded any other way, (provided that the sentence was also appropriately referenced). These exceptions were made in areas where the technical language offered no other reasonable alternatives, or similarity would be imminent.
- For example “P valves less than 0.05 were determined to be significant”, and “The XXXX program was used for statistical analysis”, “The COVID-19 pandemic is caused by the virus SARS-Cov-2 that was first discovered in Wuhan China….”. Based on this logic, we removed in its totality the methods section of all the manuscripts that we analysed. The methods section contains high similarity between papers of a similar topic, ie methodology technical language is often reused(Sun et al. 2010). Despite this, the truer indicator of plagiarism is if the copying is coming from the results section(Meo and Talha 2019). For this reason, we justified removing in its totality the methods section of each of the manuscripts that met our inclusion criteria.
- If the use of a conjunctive adverb was being used and was the only part of the report not highlighted, we did not include that word(s) in the total number of words in each sentence. Conjunctive adverbs examples include; however, furthermore, moreover, additionally, as well as repetitive statements such as “in our study” and “in their study”. We justified the removal of these words from the total word count in a sentence as this can be done to conceal verbatim text copying to avoid detection by known plagiarism detection tools or utilized by “paraphrasing” tools which change the sequence of words in a sentence or add words to avoid plagiarism.

The final “Optimized Manuscript” contained only the abstract, introduction, results, and discussion/conclusion.

Finally, an initial pilot study was conducted using n=3 papers from each quartile and examined by each of the 4 authors independently to confirm inter-rater agreement with the prescribed methods at identifying plagiarism; no differences were found between the judges.

## Statistics

Descriptive statistics were reported as percentages, frequencies, average and standard deviations. Two sample comparisons were made using t-test for normally distributed data. The optimal cut-off of the Turnitin Similarity score was explored using ROC analysis. **All statistical analysis was performed using MedCalc for Windows version 19.1 ((MedCalc Software, Ostend, Belgium)**. *p* values <0.05 were considered statistically significant.

## Results

We examined a total number of 310 articles, of which 176 were classified as original research and 134 were reviews. Of these publications, 183 were published in the year 2021 and 127 were published in 2020 (see table 2).

**Table 2:** Summary Statistics for Turnitin Similarity Report.

|  | **Quartile 1** | **Quartile 2** | **Quartile 3** | **Quartile 4** |
| --- | --- | --- | --- | --- |
| Number of papers examined | 79 | 99 | 65 | 67 |
| Average Number of Authors | 14.82±12.38 | 7.82±4.76 | 7.0±4.59 | 4.99±3.4 |
| Median Number of Authors | 10 | 7 | 6 | 4 |
| Similarity report (avg ± std) | **6.9%±5.66** | **9.89%±8.81** | **7.31%±8.28** | **8.76%±7.81** |
| Presence of plagiarism | **36.7%**<br>**(n=29)** | **52.5%**<br>**(n=52)** | **20%**<br>**(n=13)** | **52.2%**<br>**(n=35)** |
| Average # of copied sentences | 0.95±2.08 | 2.89±5.71 | 2.32±10.92 | 2.79±5.0 |
| Redundancy | 3.8%<br>(n=3) | 19.2%<br>(n=19) | 4.6%<br>(n=3) | 8.9%<br>(n=6) |
| Average Time Spent in analysis (min) | 2.44±2.09 | 2.55±2.84 | 2.25±3.8 | 2.51±3.67 |
| Far Out Reports (based on Tukey Test) | 1<br>32% | 4<br>32%, 34%, 36%,<br>61% | 3<br>33%, 33%, 40% | 0 |

The presence of PLAGIARISM, in accordance with our described methods at identifying these incidents was **41.6% (**n=129). Turnitin similarity reports revealed an average similarity of **8.35±7.84% among all analysed papers**. The lowest value of any similarity report was 0%, and the highest was 60%. Plagiarism was identified in 35% of reviews (n=47), and 46.6% of original research (n=82).

The average number of copied sentences overall the papers examined was **2.25±6.49**, based on our methods. Among those papers identified to have plagiarism, the average number of copied sentences among them was **5.42±9.18** sentences (lowest value = 1, highest value = 85 sentences).

A negative correlation was found between the year of publication and the presence of plagiarism, r=-0.122 (p=0.032 95%CI= -0.230 to -0.011). Among the papers published in 2020, 48.81% had plagiarism (n=62), while in the 2021 publications plagiarism was discovered in 36.61% (n=67).

The average number of authors on the publications were 8.82±8.13, with the lowest value being 1 and the highest being 51 authors. A negative correlation was found between the number of authors and the presence of plagiarism r=-0.1465 (P= 0.0098, 95% CI -0.2537 to -0.03565). The median number of authors was 6 (95% CI 6.0-7.0).

Based on the country of origin of the corresponding author, a publication was further determined as “English” if belonging to a core anglosphere country (USA, UK, Canada, Australia, Ireland, English speaking Caribbeans). Based on this definition 27.74% of the publications came out of core anglosphere countries (n=86), and 72.26% came from elsewhere (n=224). A significant negative correlation was found between papers published in core anglosphere countries and the presence of plagiarism, r=-0.172 (p=0.0023, 95% CI -0.278 to -0.062). See Figure 1.

**Figure 1:**
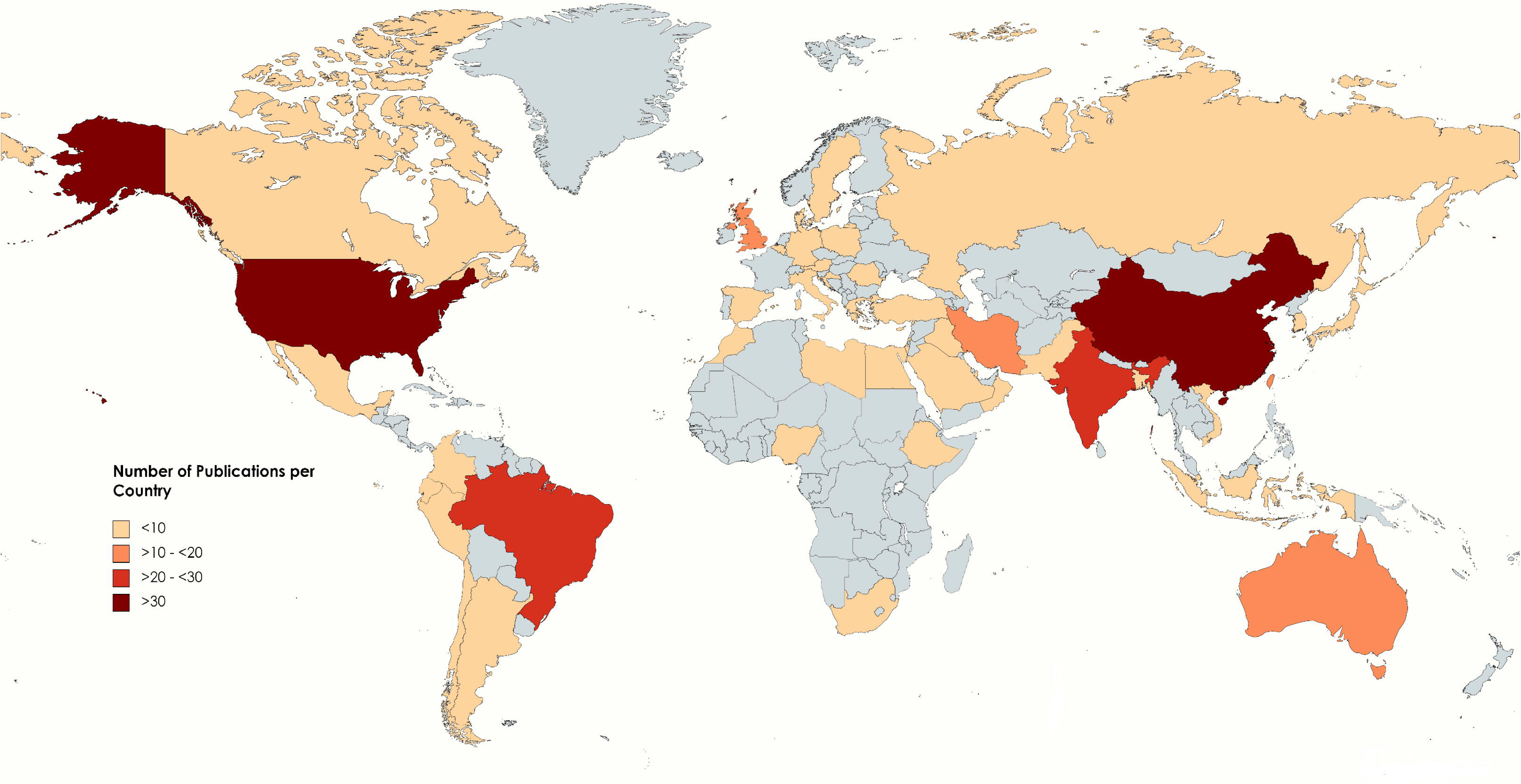
Countries of Examined Papers.

The presence of self plagiarism/redundancy was identified in 31 publications, with self plagiarism being defined as any of the authors alternative publications as the *main source* of similarity on the TURNITIN similarity report.

The average time spent analyzing the optimized reports was 2.45±3.09 minutes. The average time spent analysing the plagiarized reports was 4.53±3.80 minutes, while the average time spent on the non-plagiarized papers was 0.96±0.88 minutes.

The area with the most copying was the discussion section of the manuscripts (n=85), with the average number of copied sentences in that section being 6.25±10.16. The area with the second most number of copied sentences was the introduction with 1.6±1.47 (see figure 2).

**Figure 2:**
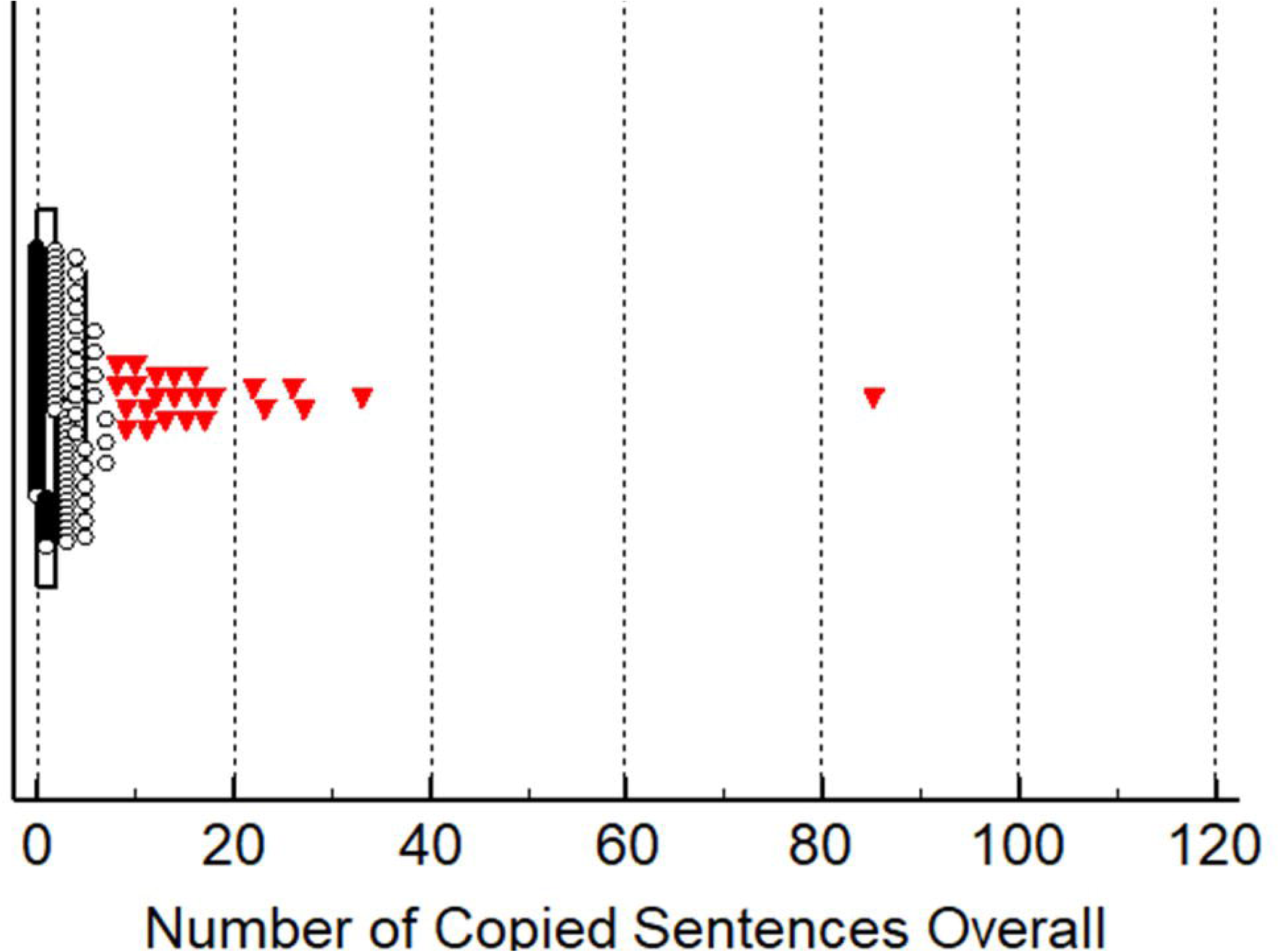
Number of Copied Sentences.

ROC analysis revealed an area under the curve (AUC) 0.828 (95% CI 0.781 to 0.868, P <0.0001) with an optimal criterion of 6% (specificity being 72.38 (95% CI 65.3 to 78.7) and sensitivity being 81.40 (95 CI 73.6 to 87.7)) (see figure 3). Based on the “accepted” similarity levels for journals – when we observe at criterion 15% sensitivity becomes **27.91** (95% CI 20.4 to 36.5) and specificity becomes 97.24 (95% CI 93.7 to 99.1). At a similarity of 18% or greater specificity remains at 99.45 (95% CI 97.0 to 100.0) and reaches 100 at similarity exceeding 35%. In other words, the 15% cut off used by some journals as an acceptable level of similarity is 97.24% specific at determining which papers *do not* have plagiarism. However, to accurately capture those publications containing plagiarism despite a similarity report of less than 15%, at Youdens criteria of 6% cut off, sensitivity is only 81.4%. In other words, a text-matching similarity-checking tool such as TURNITIN for reports less than 15% which also contains plagiarism, the tool cannot be relied on alone. Only at a criterion of 4% does sensitivity reach 90.7% (see table 3).

**Figure 3:**
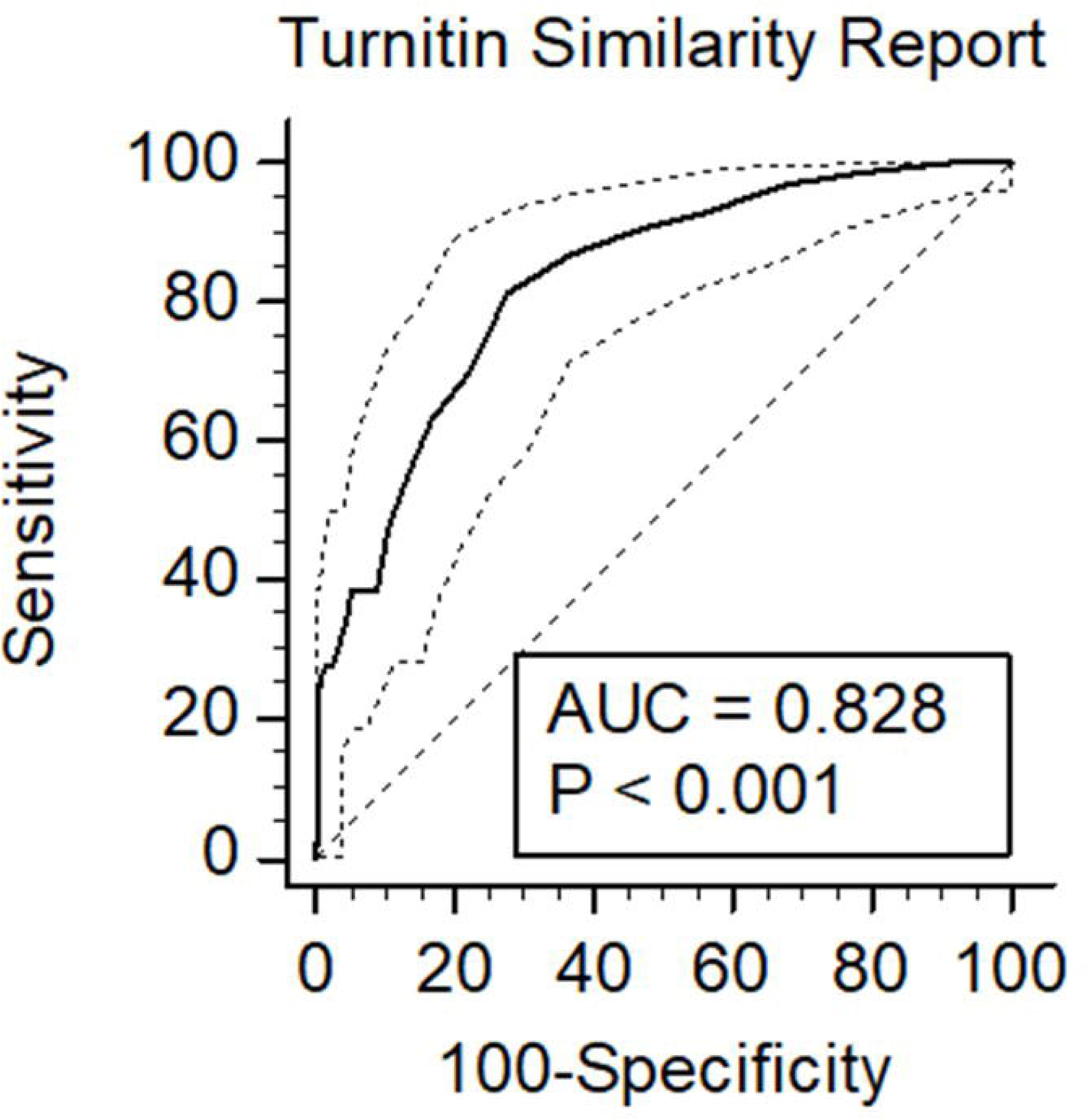
ROC curve and AUC.

**Table 3:** Range of Specificity and Sensitivity at different Criterion for Turnitin based on Our Results

| Criterion | Sensitivity | 95% CI | Specificity | 95% CI |
| --- | --- | --- | --- | --- |
| $\geq 0$ | 100.00 | 97.2 - 100.0 | 0.00 | 0.0 - 2.0 |
| $> 0$ | 100.00 | 97.2 - 100.0 | 8.29 | 4.7 - 13.3 |
| $> 1$ | 98.45 | 94.5 - 99.8 | 22.10 | 16.3 - 28.9 |
| $> 2$ | 96.90 | 92.3 - 99.1 | 32.04 | 25.3 - 39.4 |
| $> 3$ | 93.02 | 87.2 - 96.8 | 43.65 | 36.3 - 51.2 |
| $> 4$ | 90.70 | 84.3 - 95.1 | 53.04 | 45.5 - 60.5 |
| $> 5$ | 86.82 | 79.7 - 92.1 | 62.98 | 55.5 - 70.0 |
| **$> 6$** | 81.40 | 73.6 - 87.7 | 72.38 | 65.3 - 78.7 |
| $> 7$ | 69.77 | 61.1 - 77.5 | 77.90 | 71.1 - 83.7 |
| $> 8$ | 63.57 | 54.6 - 71.9 | 82.87 | 76.6 - 88.1 |
| $> 9$ | 55.04 | 46.0 - 63.8 | 86.74 | 80.9 - 91.3 |
| $> 10$ | 47.29 | 38.4 - 56.3 | 89.50 | 84.1 - 93.6 |
| $> 11$ | 38.76 | 30.3 - 47.7 | 91.16 | 86.0 - 94.9 |
| $> 12$ | 38.76 | 30.3 - 47.7 | 94.48 | 90.1 - 97.3 |
| $> 13$ | 34.11 | 26.0 - 43.0 | 95.58 | 91.5 - 98.1 |
| $> 14$ | 30.23 | 22.5 - 38.9 | 96.69 | 92.9 - 98.8 |
| **$> 15$** | 27.91 | 20.4 - 36.5 | 97.24 | 93.7 - 99.1 |
| $> 16$ | 27.91 | 20.4 - 36.5 | 98.34 | 95.2 - 99.7 |
| $> 17$ | 26.36 | 19.0 - 34.8 | 98.90 | 96.1 - 99.9 |
| $> 18$ | 23.26 | 16.3 - 31.5 | 99.45 | 97.0 - 100.0 |

This stresses the true emphasis of this research which is 1) there is high level of plagiarism in papers published on the COVID19 topic in infection related journals (41.6%), and 2) that manual curation/human judge is needed even for those papers which fall below the 15% cut off to identify the true presence of plagiarism. While a similarity report of 6% or less is not impossible (n=269, 86.77%), it is logistically difficult given contested areas of similarity such as the methods section which have re-cycled language especially among papers of a similar speciality (keeping in mind our analysis was conducted on optimized manuscripts where the methods section as a whole was removed from all manuscripts to gauge a truer sense of the similarity). Plagiarism was identified in 35.67% of the papers we examined where similarity report was 15% or less (n=93), or 31% of the total number of examined papers. Therefore, a third of the papers we examined contained plagiarism that would not have otherwise been captured without manual curation. Of those papers that exceeded the 15% cut off (n=41) – 87.8% contained plagiarism (n=36); only 11% of the total number of papers examined. In otherwords, 75% of the plagiarism containing papers belong to a threshold of 15% or less. This supports the position that the majority of plagiarism occurring is in fact below the 15% cut off (see figure 4). We examined papers from all 4 quartiles (Q1=79, Q2=99, Q3=65, Q4=67). Using Shapiro-Wilk test for normal distribution revealed normal distributions between the number of papers analysed among the quartiles (P=0.318, accept normality).

**Figure 4:**
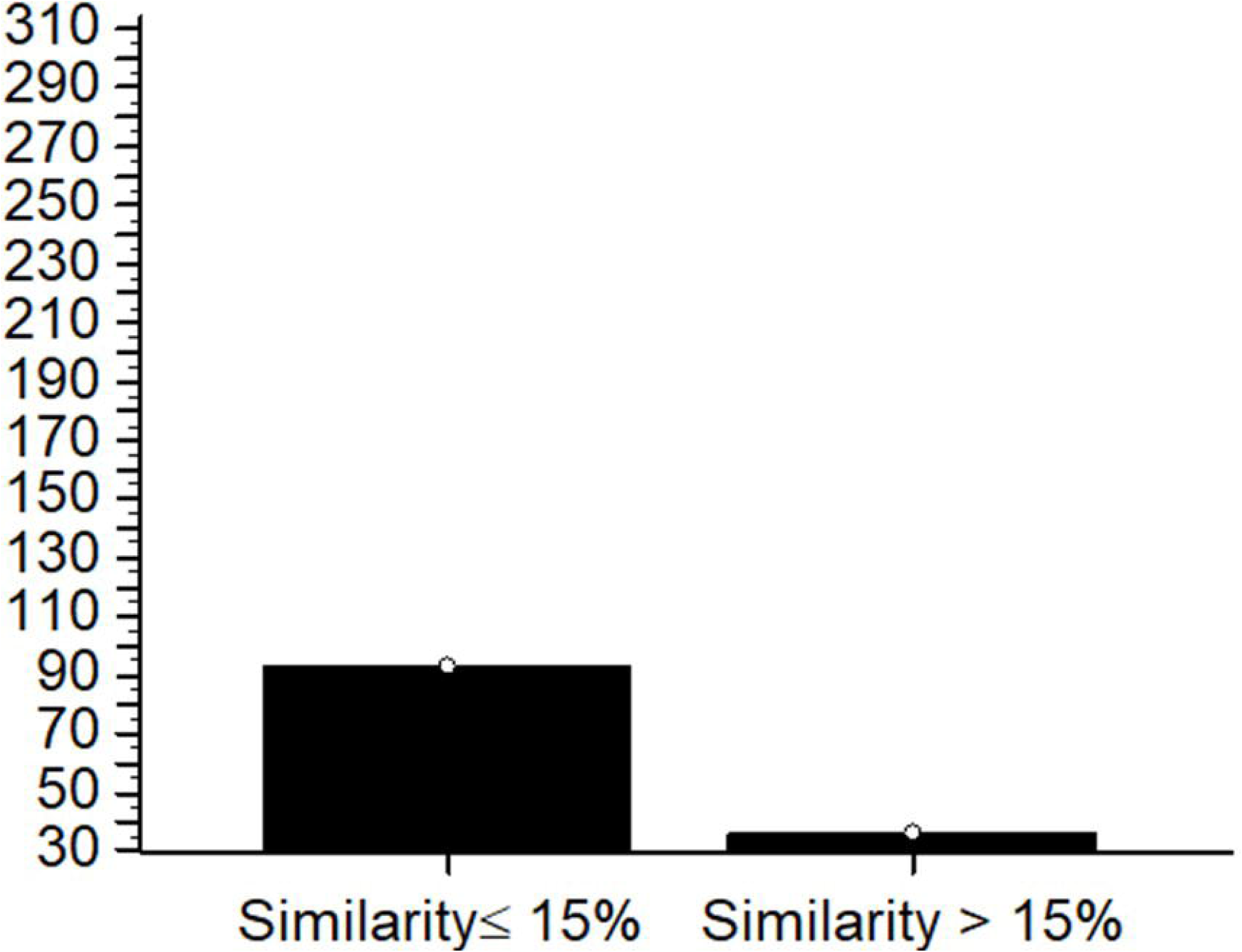
Plagiarised papers more or less than 15%.

No significant difference was found with regards to the presence of plagiarism among the quartiles (high impact = Q&Q2, versus low impact = Q3&Q4), P=0.1071.

Presence of plagiarism and similarity report results between the quartiles are detailed in table 2. Similarity reports were highest in percentage in Quartile 2, and the least in Quartile 1; **9.89%±8.81** and **6.9%±5.66** respectively (see figure 5). Number of far out similarity reports were the most in Quartile 2 (with the highest recorded similarity percentage being 61%), while Quartile 4 had no publications of serious concern. Based on the results of Tukey test for far out variables, we used this to determine which of these papers required the attention of the Editor in Chief of the respective journal to be informed of our results. Quartile 2 showed 52% plagiarism while Quartile 3 had the least with 30%. Redundancy or self plagiarism was highest in Quartile 2 (19.2%) and least in Quartile 1 (3.8%).

**Figure 5:**
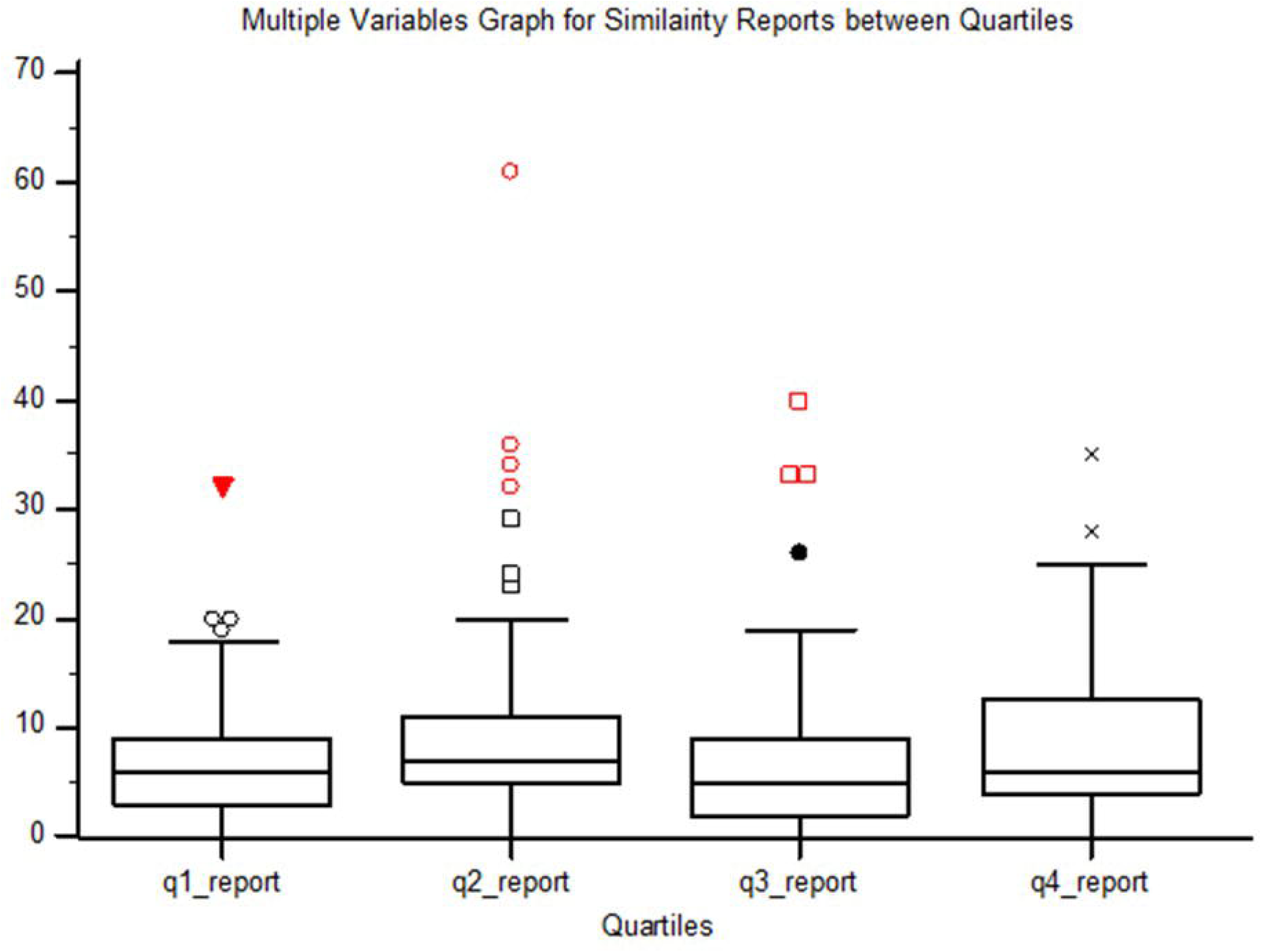
Similarity reports for all quartiles.

Lastly, while most journals we examined had some mention addressing their stance on plagiarism or their use of similarity checking tools, we wish to note that the majority of these journals rarely had these notices under a heading of their own and were often near the end of the submission guidelines. We recommend that the stance of the journal to be clearly stated under its own heading and placed earlier on in the submission guidelines to stress the importance of this issue to future authors.

## Discussion

Plagiarism is defined as adopting the work or ideas of another person as one’s own. It is a fraudulent act and considered a very serious crime in the academic setting. There are many factors which can explain, but do not excuse plagiarism. These can include academic pressure to publish, number of authors, inexperience of the authors, age of the authors, country of origin, length of the manuscript, type of the manuscript, poor citation skills, poor guidance, and much more(Debnath 2016).

We encountered two main types of plagiarism in the papers we examined. The first is known as verbatim plagiarism which is a copy-paste type. It is important to note, the direct copy and pasting another works exact text with a reference is still plagiarism. The other type of plagiarism we encountered is known as self plagiarism. Self plagiarism is the republication of one’s previous work, it is seen as a type of redundancy and so may be referred to as self-redundancy(Burdine et al. 2019).

We examined 310 consecutively submitted COVID19 papers to infection journals among 4 quartiles to determine the presence of plagiarism. We found evidence of plagiarism in 41.6% of the examined papers (n=129). We incorporated the use of the similarity checker Turnitin program as well as manual examination of the reports to determine if plagiarism existed within the text. The Turnitin program is an internet based tool developed in the year 1997 by iParadigms LLC(SA and Talha 2019). The Turnitin program is quite popular within academic institutions as a similarity checker. The Turnitin program, and much other programs like it, check the submitted work against a database of other resources such as other publications, books, journals, etc. What is yielded is a text-match result, where a highlighted section on the report denotes its presence elsewhere in the database.

There are issues to using technology designed to detect similarity as a way to also detect plagiarism. While high similarity between texts can likely suggest plagiarism is at hand, a low value does not exclude it. In our study, at a criterion of 15% similarity, the tool was only 27.91% sensitive at identifying plagiarism. On the other hand, specificity was 97.24%. Sensitivity barely reached 90% at a criterion of 4% cut off, which is logistically very difficult to achieve keeping in mind that our analysis was conducted on manuscripts without a methods section to eliminate this contested area of recycled language. For this reason, we suggest that any journal employing such text-matching software to also supplement their results with a manual judge to re-examine the similarity report for true plagiarism.

There are several blind spots within the publishing process that may miss the presence of plagiarism. As mentioned, relying solely on a text-match report without manual curation might lead to missed presence of plagiarism. Within the submission process, journals often employ such text-matching program on a manuscript before it reaches the hands of reviewers or editors. If a high similarity is observed, an immediate rejection is often sent to the authors or a request to address the issue and then resubmit. The threshold that tends to denote whether a manuscript can progress to the review process is typically less than 15% and varies per journal and speciality. The blind-spot is as follows; this 15% within the manuscript can still be all copied text, yet because it meets the journals own threshold for text-matching it bypasses and progresses in the review process. Another area of concern lies in response to reviewers where a manuscript is returned to the authors for further revisions that may have inadvertently introduced added similarity or even plagiarism to the text; it is unclear if journals employ a second re-check for the manuscripts after a response to the reviewers to avoid this.

In the context of COVID-19, drastic changes were made to the publishing framework in order to swiftly share vital information to the public and scientists regarding the urgent nature of this pandemic. Some of these changes include fast-tracked peer-review, and open access publication for all COVID-19 related papers. Pre-print services have been established which bypass the review process altogether. These changes call for intensified scrutiny for all publications(Dinis-Oliveira 2020).

Our study is the first study to our knowledge to address the presence of plagiarism in COVID19 related papers in a cross-sectional way among varying quartiles and may inspire further scrutiny in other areas or specialities that saw an influx in papers on the COVID19 topic such as pulmonology or intensive care. We found that plagiarism was highest among papers published in the year 2020, and from papers coming out of non-Anglosphere countries. We did not observe a higher incidence of plagiarism where more authors were on a paper. Among the plagiarised papers, 75% were coming from reports with 15% or less similarity which strengthens the point that this is a blind spot in the process that must be addressed. Ultimately, our results suggest that similarity/text matching tool like Turnitin can be used only to confirm the lack of plagiarism (at cut offs like 15%) but cannot be relied on alone to confirm the presence *of* plagiarism especially with lower cut off criteria. This problem can be easily remedied by the inclusion of a human judge in the process. The average time to analyse papers using the described methods was 2.45±3.09 minutes - this is a very minimal amount of time that can quickly address this problem and avoid future questions into the integrity of a published work, or in the worst-case scenario, a retraction. Plagiarism is one of the leading causes of retraction of academic publications(Campos-Varela and Ruano-Raviña 2019).

In our study, papers from quartile 2 showed the higher similarity reports, events of plagiarism, and number of copied sentences. We attribute this to the fact that quartile 2 had the most papers published out of non-core Anglosphere countries, 94% (n=93 papers).

Few other studies have examined plagiarism among papers within speciality journals. In a study by Higgins et al which examined plagiarism in 399 submitted manuscripts in a major speciality genetics journal, plagiarism was found in 17% of the analysed articles(Higgins et al. 2016). Similar to our findings, they found that plagiarism was highest in countries where English was not the official language. They spent an average time of 5.9 minutes to analyse plagiarised reports while the non-plagiarised papers took 1 minute to assess. Similarly, we discovered that plagiarised papers took an average time of 4.53±3.80 minutes to analyse while non-plagiarised papers took an average of 0.96±0.88 minutes. Higgins et al identified self plagiarism in 33 of the plagiarized reports or 53%, while in our study we observed a lower percentage of 25.58% (n=33) and this may be attributed to the relative novelty of the COVID19 topic at this time(Higgins et al. 2016).

A study by Baskaran et al examined the events of plagiarism in a major speciality andrology journal using two tools; Turnitin or iThenticate(Baskaran et al. 2019). Their Turnitin report revealed an average similarity of 8.66%±8.62. A higher mean similarity report was given by the Turnitin program, and this may be due to the larger database against which it compares the submitted work (>70 billion publications)(Baskaran et al. 2019). Unlike Basakaran et al who observed higher similarity report for reviews, in our study similarity report averages were higher in original papers compared to reviews (8.92%±9.04 versus 7.5%±5.8). Again, we hypothesize that the relative novelty of the COVID19 topic may have caused this difference, especially given that reviews were likely being extrapolated and published based on previous models of disease before original research data was available and the development of clinical trials treatment protocols and outcomes.

A study by Rohwer et al examined events of plagiarism among African Medical Journals using Turnitin and discovered 17% of papers had evidence for plagiarism(Rohwer et al. 2018). The most problematic areas were the introduction and discussion. Likewise, in our study the area with the most copied sentences was the discussion section of the manuscript.

One of the major factors believed to have influenced the rise in plagiarism seen in the past decade is the mounting academic pressure to get published(Baskaran et al. 2019). No doubt, the novelty and the worldwide focus on COVID19 has exacerbated this(Dinis-Oliveira 2020). The pressure to publish in a short period of time and be the first to speak on which treatment is showing promise, may be a risk factor for misconduct such as plagiarism or fabrication of results. The COVID19 legacy on academic integrity has yet to be scoped to its full extent, and is an important area for future research. Several alarms have been raised regarding preprint COVID19 servers which identified that a meager 5.7% of all preprints were able to meet the standards of rigorous peer review process and get published(Añazco et al. 2021). Retractions for COVID19 papers are at an all time high, with a recorded 238 retractions to date based on RetractionWatch. Intellectual dishonesty has riddled claims of miracle cures like hydroxychloroquine and ivermectin which nonetheless have taken the world by storm. These examples of are the tip of the ice-berg with COVID19 related publications and the evidence against the integrity of the literature is pointing towards substantial issues and the need to recoup and address these problems.

Plagiarism is a considered a serious offence in academia and amounts to dishonesty and a breach of ethics. Methods to prevent plagiarism include proper citation of sources, avoiding direct quotation of large parts of copied text without explicit permission from publishers, use of quotations as needed but not excessively, as well as restating the text of interest in one’s own words with proper citing of the referenced materials(Wiwanitkit 2013; Dhammi and Ul Haq 2016). Once plagiarism is detected in published work, editors may take action by directly addressing the authors and or the institutions they are associated with in order to further investigate or take punitive actions. Penalties for plagiarism can include disciplinary action, retraction of the published work, and even criminal prosecution(Kumar et al. 2014). Plagiarism can be addressed at the journal level by stressing their stance against it in journal policy or submission guidelines(Debnath and Cariappa 2018), with links to plagiarism detection tools as well using these tools themselves at two points in time – upon submission and right before publication to catch added similarity during the review process (although this cannot be used as a crutch and must also be paired with human judgement).

## Conclusions

Plagiarism is a serious unethical issue in academic science. Our results revealed the presence of plagiarism among COVID19 related publications from a variety of journals and quartiles. While the use of text-matching tools is effective at confirming the presence of plagiarism at the usual cut off/standards of similarity for most journals, it is ineffective at identifying plagiarism when similarity reports fall below that cut-off. Our results revealed that the majority of plagiarism was happening in those papers with ≤ 15% similarity. Purely relying on text-matching or plagiarism tools is not enough due to this blind spot, the solution for which requires the use of a human judge. Another blind spot may occur amid the peer review process, if when a paper is sent back for revisions, those revisions may have introduced some added similarity or plagiarism (as it is unclear if journals recheck immediately before publishing). Finally, the average time spent in analyzing the manuscripts was 2.45±3.09 minutes. This is a very minimal amount of time to weed out problematic papers or alert the authors of the issue in order to maintain a high ethical standard of the papers being published in a journal and avoid misconduct and retractions.

We recommend that authors employ ethical writing standards in their work and use resources available to them to identify areas of concern in their manuscript, and for journals to utilize plagiarism checking tools along with manual curation of the reports, to ensure that a truly original paper has entered the scientific literature. It is of paramount importance to maintain due diligence with quality of work being published in this era, and that credit is properly given to the work of others, in order to maintain our standard of ethics and honesty in these trying times.

## Limitations

There are some limitations to our study. First, we only examined papers indexed on SCOPUS infection journals list, therefore our results cannot be extrapolated to represent the status of all infection journals as we did not consider those journals. Secondly, we set the Turnitin settings to examine only through journals, periodicals, and publications. We did not include other repositories such as student and institution papers. Inclusion of these repositories may have increased the overall similarity reports.

## Data Availability

All data produced in the present study are available upon reasonable request to the authors

## Ethics Declarations

### Author Contributions

RM & EM devised the study question and idea. RM, EM, AM, BM participated in data collection, analysis and writing of the manuscript.

### Ethical Approval

This study was conducted on publicly available information using a commercially available program. It was exempt from review and approval.

